# Somatic instability of the *C9orf72* repeat expansion in blood differs between patients with amyotrophic lateral sclerosis or frontotemporal dementia

**DOI:** 10.64898/2026.10.06.26364580

**Authors:** Mathieu Barbier, Mounir Ichalalen, Maria-Del-Mar Amador, Julie F. H. De Houwer, EunRan Suh, François-Xavier Lejeune, Théophile Bieth, Cécile Reyes, Emilie Henry, Allyson E. Hinks, Philippe Martin-Hardy, Daisy Rinaldi, Lara Araktingi, Harro Seelaar, Carole Roué-Jagot, The French clinical and genetic research network on FTLD / FTLD-ALS PrevDemALS and STRATALS,, Julien Lagarde, Marie Sarazin, François Salachas, Gaëlle Bruneteau, Marc Teichmann, Richard Levy, Elsa Bernard, Laura Donker Kaat, Viviana Van Deerlin, Georg Haase, John van Swieten, Audrey Rapinat, David Gentien, Isabelle Le Ber

**Author notes:** Corresponding author: Mathieu Barbier.

## Abstract

Individuals carrying a G4C2 repeat expansion in *C9orf72* can develop amyotrophic lateral sclerosis (ALS) and/or frontotemporal dementia (FTD), at various and unpredictable ages. The contribution of the expansion to this extreme clinical variability remained a matter of debate. In this work, we validated the use of optical genome mapping to efficiently size this expansion from blood and applied this method to a cohort of 54 repeat expansion carriers including patients with FTD (n = 23), ALS (n = 22) and presymptomatic individuals (n = 9). Despite the high variability in expansion profiles, distinct groups can be inferred. Although consensus repeat profiles were similar between FTD and ALS patients, relationships between repeat number, indexes of repeat instability, and the age at sampling differed between clinical conditions. An age at sampling-dependent increase of repeat instability in blood was observed in presymptomatic carriers and FTD patients (p = 0.01 and p = 0.04 respectively), in contrast to patients with ALS (p = 0.9). The proportion of shorter repeats ([500-1500 repeats]) increased with the age at sampling in ALS patients but decreased in presymptomatic and FTD cohorts, while the proportion of larger repeats (>2000 G4C2), in particular those exceeding 4000 repeats, increased along with the age at sampling in presymptomatic carriers and FTD cases (p <1.10^−e4^ and p = 0.006 respectively) conversely to patients with ALS (p = 0.98). In conclusion, this work strongly supports different dynamics of repeat instability of the *C9orf72* pathogenic expansion in blood linked to the clinical evolution. These findings provide new insights into the relationship between expansion profiles and clinical variability.

## Introduction

The G4C2 expansion in the first intron of the gene *C9orf72* is the major cause of inherited frontotemporal dementia (FTD) and amyotrophic lateral sclerosis (ALS)^1,2^. Clinical variability is a hallmark of *C9orf72*-related disease illustrated by the occurrence of ALS and/or FTD. In addition, the age at onset spans from the third decade of life to, in some carriers, over 90 years. Patients carry from hundreds to thousands of repeats. Consequently, the search for correlations between the variability of clinical symptoms and repeat number emerged early after the discovery of this pathological expansion as in other repeat expansion disorders, sometimes yielding conflicting results from Southern blot-based studies^3–9^. More recently, this pathological expansion, previously unsequenceable by conventional or short-read methods, has been successfully sequenced in its entirety using long-read sequencing approaches (LRS)^10–13^. Interestingly, LRS-based studies depicted a partial overlap between repeat estimates and some clinicopathological associations (notably with survival after onset), showed that the repeat content was almost pure, and allowed the analysis of methylation levels^11–13^. This illustrates how new tools can bring additional knowledge in the field. Improvements of LRS approaches now allow the detection of long expansions^12^. However, LRS still substantially underestimates the mean number of G4C2 repeats compared with Southern blotting^11–13^. A bias toward the sequencing of the wild-type allele as well as shorter expansions could lead to truncated allelic distributions of different repeat numbers affecting large repeats, hampering a global view of the somatic variability of the *C9orf72* expansion.

Optical Genome Mapping (OGM) has emerged as a promising technology for the detection of pathological repeat expansions, including *C9orf72*^14^. To date, the efficacy of OGM to size the *C9orf72* expansion has never been thoroughly tested in a large cohort. Here we validate the use of OGM to quantify the repeat number in molecules carrying *C9orf72* pathological expansions leading to a more comprehensive expansion profiling, with an improved detection of very large expansions. Furthermore, we applied this technology to a cohort of 54 *C9orf72* repeat expansion carriers with variable clinical conditions (presymptomatic, ALS or FTD symptoms at sampling) not only to examine relationships between repeat number and clinical traits in the light of this new method, but above all to consider repeat profiles and associated somatic variability to clarify the role of this expansion in clinical variability.

## METHODS

### Subjects and ethics

Forty-five patients (23 with FTD and 22 with ALS at sampling) known carriers of the pathogenic G4C2 expansion in *C9orf72* were included in this study: 29 of French origin, 13 Dutch, and 3 North American. Nine presymptomatic *C9orf72* expansion carriers (4 French and 5 North American) were also included. Detailed information about this cohort of 54 repeat expansion carriers can be found in Table 1. Genetic status of all individuals has been established for all of them by repeat-primed PCR prior to inclusion. Clinical data and blood samples were obtained after written informed consent for clinic-genetic studies was signed.

**Table 1.** Descriptive data of the cohort. Principal metrics that were obtained from the analysis of the *C9orf72* expansion by optical genome mapping are also mentioned. Data are reported as median (Q1, Q3). GGGGCC group comparisons were performed using individual subject medians. Group differences were evaluated by Kruskal-Wallis test, with ^a^pairwise comparisons between Frontotemporal Dementia (FTD) and Amyotrophic Lateral Sclerosis (ALS) assessed via two-tailed Wilcoxon rank-sum tests.

|  | FTD | ALS | Presymptomatic | *p* |
| --- | --- | --- | --- | --- |
| n | 23 | 22 | 9 |  |
| Males, n (%) | 12 (52%) | 12 (55%) | 3 (33%) |  |
| Age at sampling (y) | 65 (58, 74) | 61 (56,70) | 41 (37, 53) | 0.002; 0.21<sup>a</sup> |
| Age at onset (y) | 63 (52, 66) | 58 (54, 68) | - | 0.9 |
| Disease duration at sampling (y) | 5 (3, 9.5) | 2 (1, 3) | - | 0,0002 |
| *C9orf72 Optical Genome Mapping* |  |  |  |  |
| Molecules with expansion (n) | 74 (61, 80) | 68 (60, 81) | 66 (64, 81) | 0,9 |
| *GGGGCC repeat number* |  |  |  |  |
| Mean | 1880 (1431, 2159) | 1666 (1390, 2040) | 1863 (1669, 2074) | 0.65; 0.43<sup>a</sup> |
| Median | 1676 (1191, 1882) | 1536 (1262, 1954) | 1697 (1593, 1869) | 0.57; 0.68<sup>a</sup> |
| Standard Deviation | 1002 (754, 1300) | 912 (568, 1192) | 799 (590, 1103) | 0.45; 0.30<sup>a</sup> |
| Maximum repeat number | 6629 | 6669 | 7611 |  |

Ethic committee approvals were obtained for the French individuals (IRB: Comité de Protection des Personnes (CPP) Ile de France II for INSERM (#RBM 02-59) and CPP Sud-Est IV for BIOMOV collections (APHP #2021-A00989-32). Dutch individuals were part of the existing FTD-RisC cohort at the Erasmus MC University Medical Center which received approval from the ethical committee of Erasmus MC University Medical Centre (MEC-2009-409). Eight individuals from the University of Pennsylvania were included in the study who participated in an informed consent procedure that was approved by an Institutional Review Board (IRB #842873).

### Sizing of the C9orf72 expansion by Optical Genome Mapping

The principle of OGM applied to the *C9orf72* expansion is to perform a physical mapping of stained DNA, to analyze distances between markers, and then to extrapolate the repeat number based on the abnormal distance between two markers encompassing the *C9orf72* expansion locus compared with a consensus optical map. Fresh frozen blood samples were used to ensure sufficient quality of DNA extracted. Ultra-high molecular weight genomic DNA (UHMW gDNA) was extracted from frozen EDTA whole blood using the Bionano Genomics Prep SP Blood & Cell Culture DNA Isolation Kit following the manufacturer’s protocol (CG-00006, Rev E). DNA concentration was measured with the Qubit dsDNA Broad Range assay (Q32853). For downstream DLS-G2 labeling, gDNA concentration was adjusted to 39–150 ng/µL. Labeling of DNA was performed with the Bionano Genomics Prep Direct Label and Stain-G2 (DLS-G2) kit following the manufacturer’s protocol (CG-30553-1, Rev F). For each sample, 750 ng gDNA were brought to 19.5 µL and mixed with a master mix containing the DLE-1 labeling enzyme and the DL-Green fluorophore substrate. The reaction was incubated for 1 h at 37 °C and then stopped by adding proteinase K and incubating for 30 min at 50 °C. Labeled DNA was cleaned up on the DLS membrane to remove unincorporated DL-Green; the DNA backbone was stained with the kit DNA Stain and homogenized for 30 min. Labeled/stained DNA was quantified with the Qubit dsDNA High Sensitivity assay (Q32854) and adjusted to 4–16 ng/µL prior to Saphyr chip G3.3 loading. BNX molecule files generated by the Saphyr system were aligned using Bionano Genomics’s Align Maps (RefAligner) workflow to an in silico–digested hg38 reference. The pipeline produced the standard alignment triplet r.cmap (reference), q.cmap (query), and xmap (alignment table), which served as inputs to an in-house script to estimate GGGGCC repeat number on a per-molecule basis (script available at https://github.com/Genosfert/ogm-tandem-repeat-sizing). OGM, as well as long-read sequencing of repeat expansions, can give allelic distributions of repeat numbers, with an overlap between wild-type and expanded alleles. For each sample, a two-component Gaussian mixture model (GMM) was fitted to the distribution of expansion sizes, and the intersection point of the two Gaussian density functions, defined as the Bayesian threshold, was used to distinguish between non-expanded and expanded molecule subpopulations. A global Bayesian threshold was then estimated as the median of the individual thresholds, and its 95% confidence interval was calculated using non-parametric bootstrap resampling (2000 iterations). In the studied cohort, the resulting global threshold was 65.25 repeats [95% CI: 61.23–67.01], supporting the use of an operational and conservative threshold of 70 G4C2 repeats to classify a molecule as carrying a pathogenic expansion for downstream analyses.

### Sizing of C9orf72 expansion by Oxford Nanopore Long Read Sequencing (ONT-LRS)

Genomic DNA was extracted from frozen EDTA whole blood using the QIAGEN QIAamp DNA Mini Kit (Ref. 56304) following the manufacturer’s protocol. CRISPR/Cas9-targeted Nanopore sequencing was performed using the Cas9 Sequencing Kit (SQK-CS9109, Oxford Nanopore Technologies). Two crRNAs flanking the *C9orf72* repeat locus were designed and annealed with tracrRNA (100 µM each, IDT) at 95°C for 5 min, then cooled to room temperature to generate guide duplexes. Ribonucleoprotein (RNP) complexes were assembled by combining the guide duplexes with HiFi Cas9 nuclease in ONT Reaction Buffer and incubating for 30 min at room temperature. Genomic DNA (1.5–7 µg) was dephosphorylated using the phosphatase supplied in the kit (37°C for 10 min, followed by 80°C for 2 min). Dephosphorylated DNA was then incubated with RNP complexes together with dATP and Taq polymerase to perform Cas9 cleavage and simultaneous dA-tailing (37°C for 30 min, then 72°C for 5 min). Sequencing adapters from the SQK-CS9109 kit were ligated at 16°C for 2h 30 min using T4 DNA ligase according to ONT recommendations. Adapter-ligated fragments were purified using 0.3X AMPure XP beads with Long Fragment Buffer (LFB) to enrich for long DNA molecules and eluted in 12 µL Elution Buffer. Libraries were loaded onto R9.4.1 (FLO-MIN106) flowcells following standard ONT priming and SpotON loading procedures and sequenced on a MinION Mk1B for up to 48 h.

Basecalling was performed using Guppy v6.5.7 with the High-Accuracy (HAC) model (dna_r9.4.1_450bps_hac.cfg). Reads were aligned to the GRCh38/hg38 reference genome using Minimap2 (v2.17), and *C9orf72* G4C2 repeat numbers were quantified using STRique (v0.4.2).

### Sizing of the C9orf72 expansion by Southern blots

Blood samples from eight patients (five presymptomatic and three FTD expansion carriers) for whom the *C9orf72* repeat number was estimated using OGM were also used for *C9orf72* expansion sizing by Southern blots. Importantly, familial histories of these expansion carriers did not reveal specific enrichment for motor or cognitive symptoms in affected relatives, histories of ALS, FTD, or ALS/FTD were reported. The mode, defined as the point on the Southern blot with the highest intensity, was used as the repeat number for each individual as described previously^5^.

### Statistics

All statistical analyses were performed using R v4.3.2 (R Core Team, 2023). Between groups comparisons were evaluated by Wilcoxon rank-sum and Kruskal–Wallis tests for continuous variables, and the Fisher’s exact test was used for categorical variables, as appropriate. Unsupervised hierarchical clustering of repeat number distribution was performed using Wasserstein’s distance. Pairwise Wasserstein-1 distances between participants were calculated using the wasserstein1d function from the transport R package (v0.15-4), followed by hierarchical clustering using complete linkage. Wasserstein’s distance is well suited for comparing continuous distributions, as it is sensitive to their shape, peaks, and dispersion. A linear mixed-effect model including sex, age at sampling and condition (ALS of FTD) as fixed effects, with sample included as a random intercept, was used to assess the independent association with G4C2 repeat number. Beta, CI and unadjusted p-value are indicated. Spearman’s correlation was used for all correlations. In regression plots, fitted regression lines from linear regressions also appear to visualize the direction of the correlation. The “index of somatic instability” used in this work was calculated as described for other repeat expansion disorders^15^. This index was calculated as the absolute difference between the 90th percentile and 10^th^ percentile of the allelic distribution (expanded alleles) measured in each sample. All tests were two-sided. PRISM v10 software was also used to generate some figures.

## Results

### Detection of the C9orf72 pathogenic expansion by Optical Genome Mapping

The *C9orf72* pathological expansion was detected in all individuals, consistent with their genetic diagnosis. The number of molecules carrying the pathogenic expansion was homogeneous in the cohort and did not vary significantly across phenotypes (mean number of expanded molecules per sample = 68.50 ± 11.65; Table 1; Supplementary Figure 1A). The detection of molecules carrying pathogenic expansions was not correlated with the age at sampling (ρ = -0.05, p = 0.73). Importantly, the mean ratio of the allelic balance between wild-type and expanded molecules was equal to 0.50 ± 0.04 as expected for heterozygous carriers (Supplementary Figure 1B). This unbiased ratio did not vary across individuals’ clinical status and ages at sampling (data not shown). Standard deviation of repeat distribution was highly correlated to the mean repeat number of the expanded allele in the whole cohort (ρ = 0.64, p < 1.10^e-4^). The maximum number of G4C2 repeat number detected in a molecule was 7611. Two patients with different expansion profiles were sampled twice at 6-month and one-year intervals (Supplementary Figure 2). The expansion profile of the patient presenting a Gaussian-like distribution was replicated. The other carrier presented a more complex profile with a pseudo bi-modal distribution of repeat numbers. This pattern was also replicated. Numbers of repeats were in the same range for each replicate. Both mean and median repeat numbers were similar between men and women (p = 0.78 and p = 0.88 respectively). In line, the comparison of allelic distributions according to the sex did not reveal significant differences (p = 0.88, Supplementary Figure 3).

### Comparisons of OGM with Southern blotting and ONT long-read sequencing

A comparison of median G4C2 repeat number estimates by OGM and repeat number estimates from Southern blot, the initial gold-standard method, was performed in eight independent samples to assess the robustness of OGM and validate our informatic pipeline. Median repeat numbers calculated by OGM were strongly correlated with those obtained by Southern blots (ρ = 0.88; p = 0.0039; Figure 1A). In line, repeat numbers per individual were relatively similar between OGM and Southern blots (Figure 1B).

**Figure 1.**
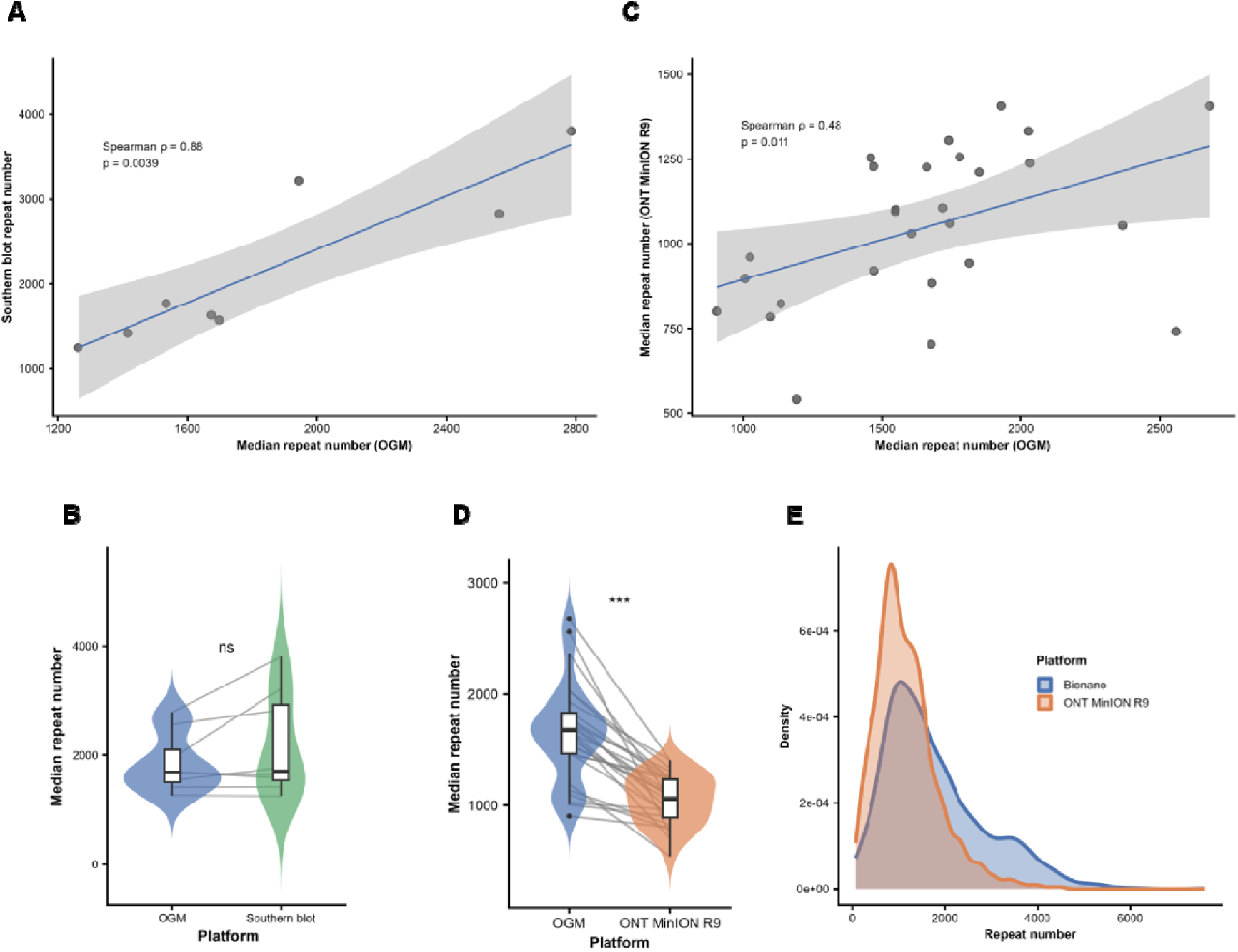
Optical Genome Mapping (OGM) to size the G4C2 pathological repeat in *C9orf72*. **A-B:** Comparison with Southern blotting. **A)** correlation between OGM and Southern blots. **B)** Matched comparison of median repeat number. **C-E**: Comparison with ONT long read sequencing (ONT-LRS). **C)** Scatterplots showing the correlation between OGM and ONT-LRS. The fitted regression line from linear regressions appears. **D)** Matched comparison of median repeat number. **E)** Allelic distribution of expanded alleles using OGM and ONT-LRS (density curves). In (A) and (C), individual measurements from both methods are plotted with fitted linear regression lines, 95% confidence intervals (shaded areas), and annotated Spearman’s ρ and p-values. In (B) and (D), comparisons between methods are displayed using violin plots and overlaid boxplots, with line segments connecting paired individual measurements.

Parallel sizing of this expansion by ONT-LRS using R9 flowcells was also performed in 27 samples to compare classical metrics and repeat expansion profiles. OGM and ONT-LRS estimates of median G4C2 repeat number were significantly correlated (ρ = 0.48; p = 0.011; Figure 1C), although median repeat numbers were significantly lower with ONT-LRS vs. OGM in paired comparisons (Figure 1D). The modes of the distributions were similar but the consensus distribution using OGM was more enriched in large expansions with more than 2000 repeats (Figure 1E). The Bland-Altman plot and analysis confirmed that ONT-LRS underestimated the repeat number by a mean of -607.6 repeats compared with OGM, and that this bias increased as the expansion became larger (Supplementary Figure 4). Thus, the technical comparison between Southern blots, OGM, and ONT-LRS revealed concordant results between OGM, Southern blots, and ONT-LRS. Data from the allelic distributions (expanded alleles) obtained by OGM were then used for subsequent analyses.

### Interindividual variability of C9orf72 repeat expansion profiles

The distribution of molecules carrying the pathogenic expansions was highly variable at the individual level, ranging from Gaussian-like distributions to more complex ones, with virtually unique profiles for each patient (Figure 2). However, the unsupervised clustering of these distributions revealed different statistical clusters, corresponding to distinct repeat profiles (Figure 3). The resulting clustering cannot clearly distinguish symptomatic from pre-symptomatic carriers nor patients with ALS or FTD. Neither sex nor age at sampling and time from onset seemed to strongly influence the clustering. However, three main clusters can be distinguished, characterized by different consensus profiles, confirming the effectiveness of the metric used for clustering (Figure 3B, Supplementary Table 1). Clade 1 (n = 23) is characterized by highly similar distributions (short Wasserstein distances), and a lower somatic instability than clusters 2 and 3 (Kruskal-Wallis test, p = 0.00016; Dunn-Holm post hoc test, Cluster 1 vs. Cluster 2: p = 0.0050; Cluster 1 vs. Cluster 3: p = 0.0038). The allelic distribution was centered around 1,000 G4C2 repeats and included a mix of ALS and FTD patients. Clade 2 (n = 20) was intermediate in terms of somatic variability. Interestingly, clade 3 included a majority of patients with FTD (8/11). The consensus distribution of clade 3 is principally characterized by larger expansions and a higher degree of somatic instability. Thus, this unsupervised clustering revealed that despite highly variable expansion profiles between carriers, these profiles can be clustered in distinct groups.

**Figure 2.**
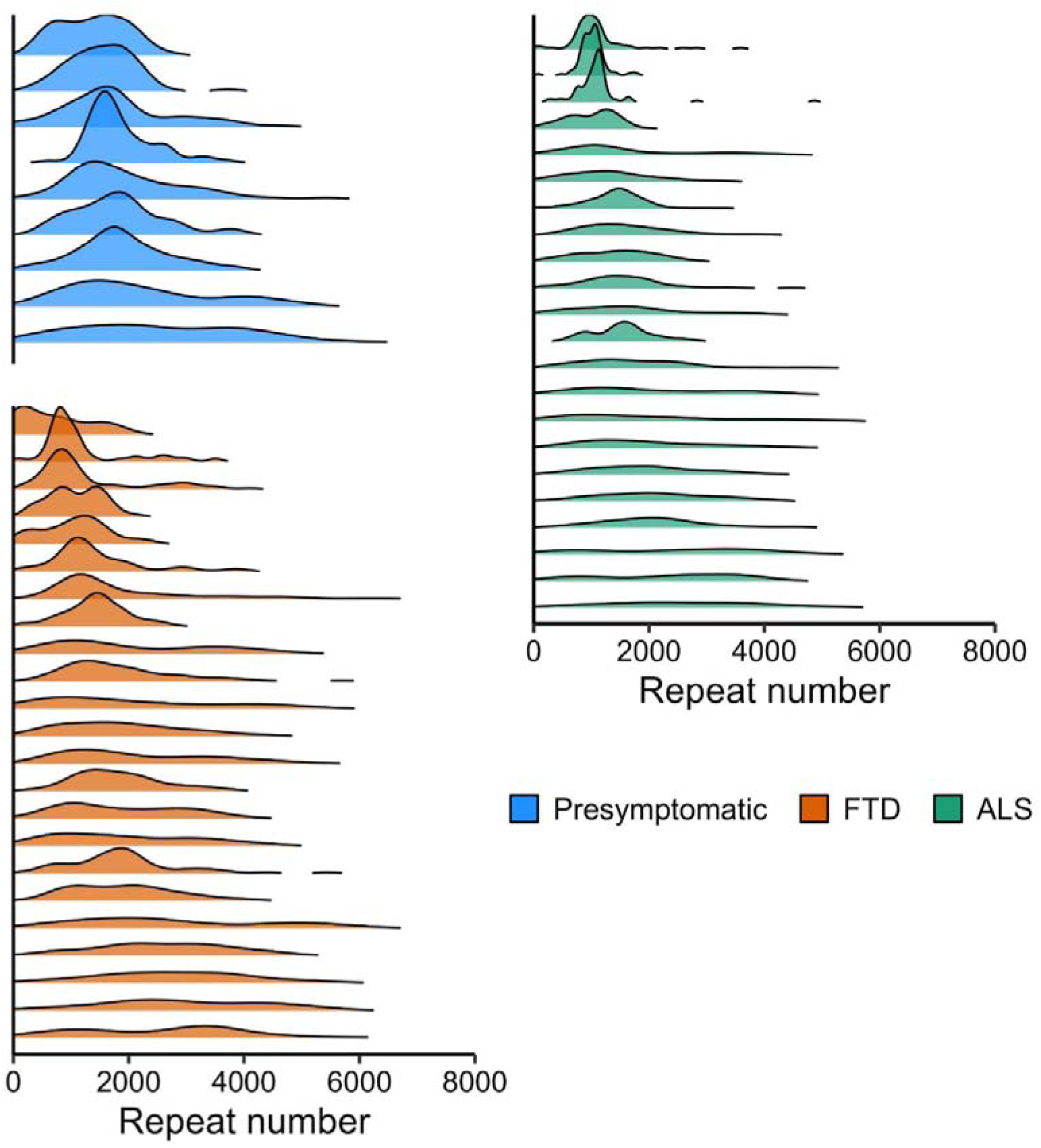
Density curves from the allelic distribution of the *C9orf72* expanded alleles for each carrier (n=54). Profiles are sorted by clinical status and increasing age at the time of sampling (bottom to top = youngest to oldest within each clinical status).

**Figure 3.**
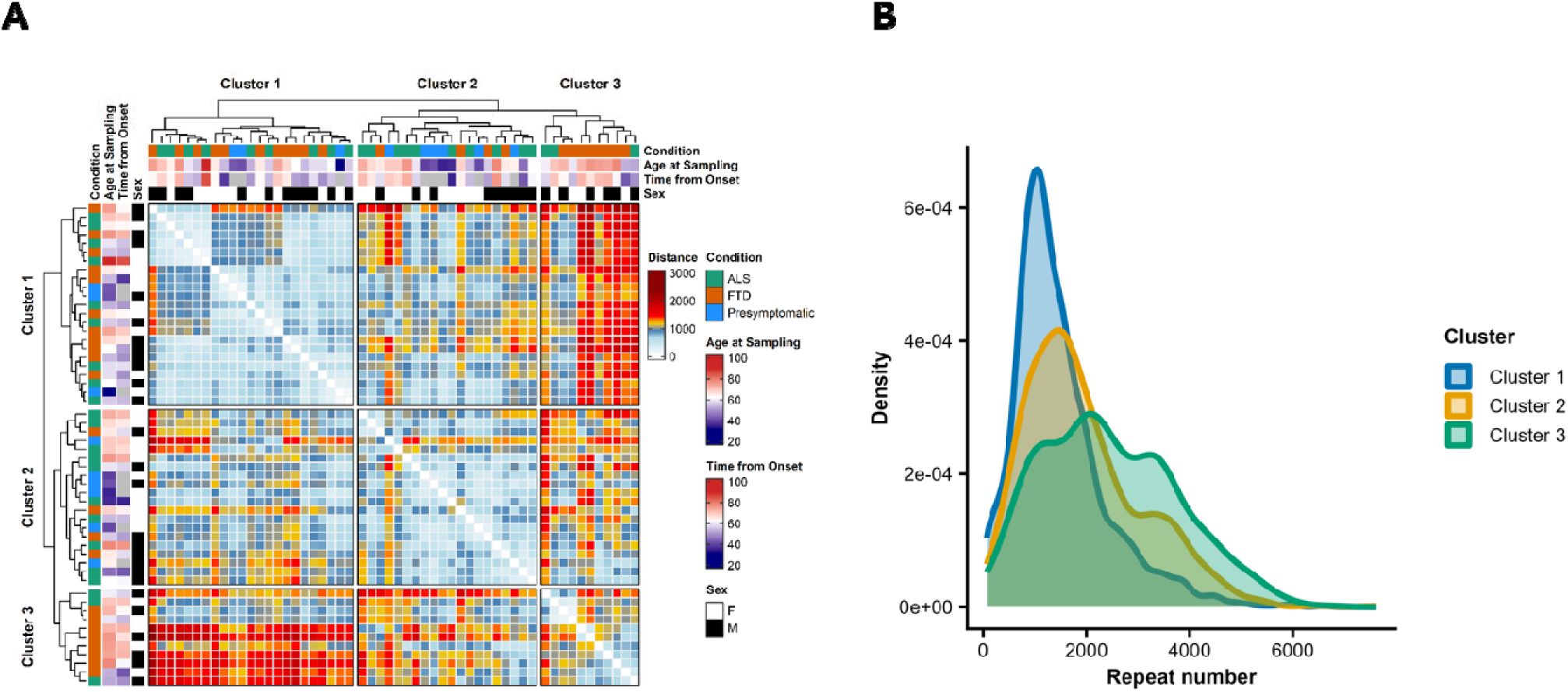
Variability of expansion profiles among *C9orf72* repeat expansion carriers. **A)** Pairwise 1-Wasserstein distances between individual repeat expansion distributions were calculated, followed by unsupervised hierarchical clustering using complete linkage, revealing three main clusters. Results are shown as heatmap (ComplexHeatmap R package v2.18.0) annotated with clinical condition (presymptomatic, ALS, or FTD), age at sampling (years), time from onset (years), and sex. **B)** Consensus expansion profiles drawn from the main clusters highlighted by the unsupervised clustering.

### Variability of repeat expansion profiles and clinical status

Cohorts of patients (with FTD or ALS at sampling) were matched for ages at sampling and sex ratio (Table 1). FTD patients tended to have a higher proportion of larger repeats (> 3000 G4C2 repeat number) compared with ALS patients while the consensus distribution drawn from ALS patients suggested a higher proportion of smaller repeat (1000-2000) (Figure 4A). This trend was confirmed by the analysis of preferential size enrichment in ALS vs. FTD (Figure 4B), although the comparison of these two consensus distributions was not significant (p = 0.69), which was also in line with the unsupervised clustering unable to discriminate FTD and ALS patients. A linear mixed-effects model incorporating age at sampling, sex, and condition (FTD or ALS) as fixed-effect predictors was used to assess the independent association of each covariate with mean G4C2 repeat number (Table 2). FTD status was associated with a non-significant increase in repeat number compared with ALS, independently of the age at sampling, whose association with repeat number was weak in this cohort combining both phenotypes. Thus, the global comparison of repeat profiles in patients did not reveal major differences between FTD and ALS, although FTD status tended to be associated with higher repeat numbers, independently of the effect of the age at sampling.

**Figure 4.**
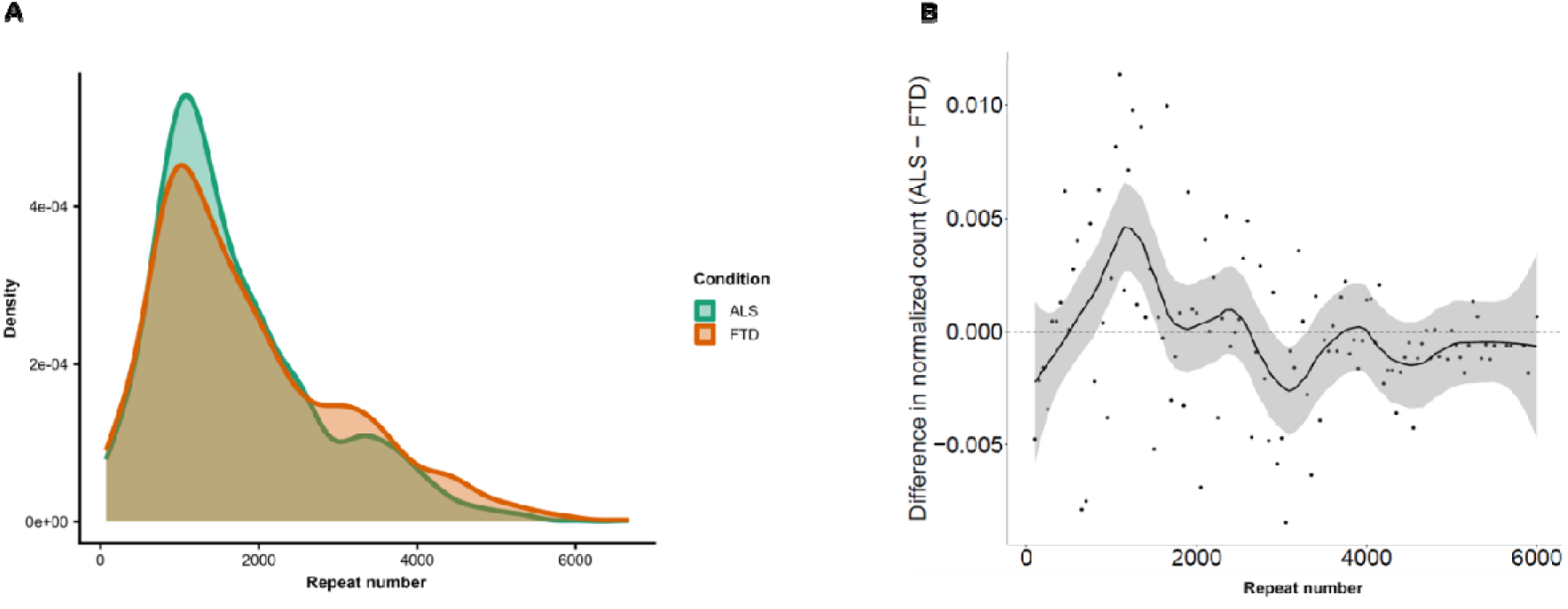
*C9orf72* expansion profiles of ALS and FTD patients. **A)** Consensus allelic distributions of expansion profiles in ALS (green) and FTD (orange) patients. **B)** Preferential size enrichment analysis: repeat number bins (50 repeats) above the dotted line are enriched in ALS cases, whereas those below the line are enriched in FTD cases.

**Table 2.** Effect of age at sampling, sex, and condition on *C9orf72* repeat number in patients with FTD or ALS at sampling. (Linear mixed-effects model regression; CI: 95% confidence interval).

| Variable |  | n | Estimate (CI) | p |
| --- | --- | --- | --- | --- |
| Age at sampling |  | 45 | 4.90 (-155.35, 165.14) | 0.95 |
| Sex | F | 21 | Reference |  |
|  | M | 24 | -78.58 (-394.24, 237.09) | 0.63 |
| Condition | ALS | 22 | Reference |  |
|  | FTD | 23 | 148.05 (-172.81, 468.91) | 0.37 |

Indeed, the age at sampling is known to influence *C9orf72* repeat number independently of disease onset^3^ making difficult the interpretation of correlations between the age at onset and clinical outcomes, as ages at sampling and onset are usually highly correlated. This was also the case in our cohort (ρ = 0.92, p < 1.10^e-4^). Consequently, we focused on the age at sampling to avoid confounding factors, but all trends described below were also observed when considering the age at onset (data not shown). Median repeat number was not correlated with the age at sampling in the whole cohort (p = 0.98), in line with the glm analysis. Surprisingly, correlations analyses considering each clinical status separately revealed distinct correlation patterns between presymptomatic carriers, FTD and ALS patients (Figure 5A). A positive correlation between the median of repeat number and age at sampling was observed in presymptomatic carriers as well as for FTD patients (ρ = 0.95, p = 6.10e^−5^ and ρ = 0.15, p = 0.50 respectively). Conversely, a negative correlation was observed in ALS patients (ρ = - 0.30, p = 0.18). Even more distinct were the differences of the somatic instability in function of the age at sampling (Figure 5B). This index of repeat instability increased significantly with the age at sampling in pre-symptomatic carriers and FTD patients (ρ = 0.78, p = 0.01 and ρ = 0.42, p = 0.045 respectively) whereas somatic instability appeared variable among ALS cases and not correlated at all with the age at sampling (ρ = -0.03, p = 0.90).

**Figure 5.**
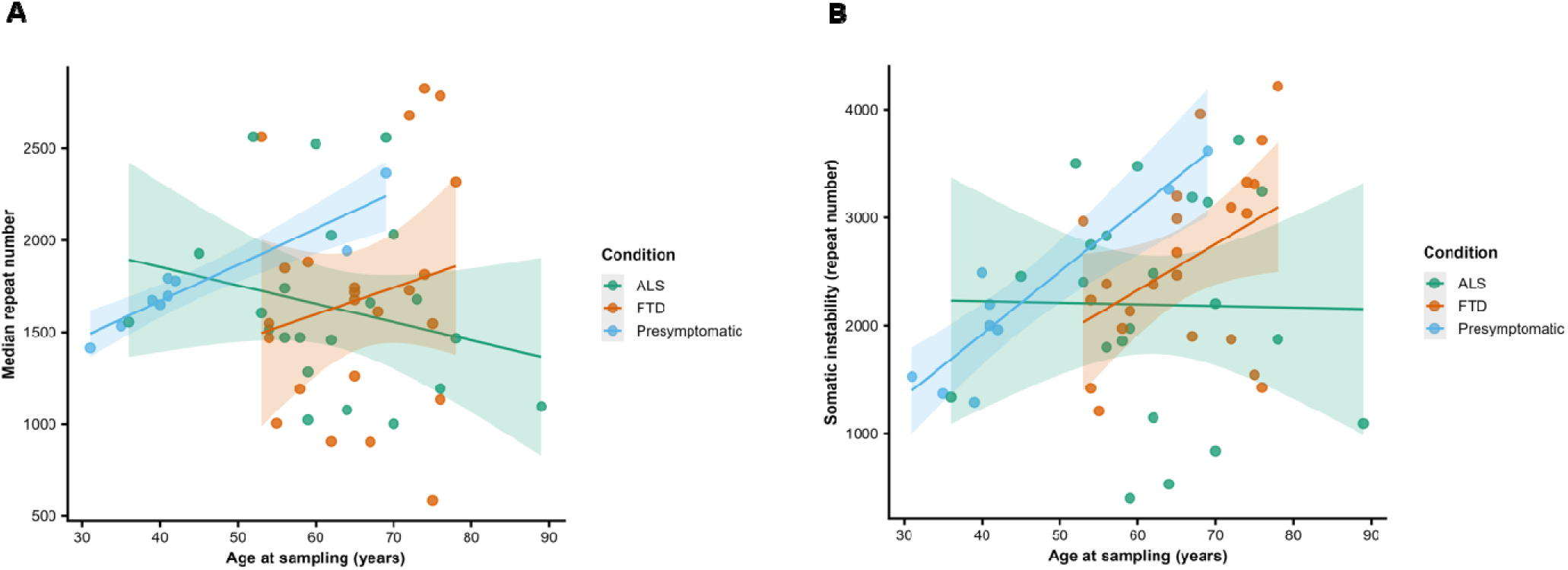
Relationships between *C9orf72* G4C2 repeat number, somatic instability, and age at sampling. **A)** Scatterplot showing the correlation between median repeat number and age at sampling in ALS (green), FTD (orange) and pre-symptomatic (blue) carriers. **B)** Somatic instability of the expansion (ΔQ90-Q10) in ALS (green), FTD (orange) and pre-symptomatic (blue) carriers as a function of the age at sampling. In (A) and (B), individual data points are plotted with fitted linear regression lines and 95% confidence intervals (shaded areas).

To better decipher these discrepancies between presymptomatic, FTD and ALS expansion carriers, the correlation between age at sampling and repeat number was assessed for each interval of repeat number (bins width of 500) (Figure 6). The only bin with similar trends of correlation between was 1500-2000, for which the proportion of corresponding repeats decreased in aged carriers, regardless of their status. This drop of proportion was in favor of smallest repeat numbers in patients with ALS between 500 and 1500 G4C2 repeats for whom the proportion increased along with the age at sampling and decreased in presymptomatic and FTD expansion carriers (Supplementary Table 2). On the other side, the proportion of large expansions from 2000 repeats onward increased along with the age at sampling for almost all repeat intervals in presymptomatic and FTD and not in ALS cases for whom the proportion of these bins was stable or even slightly decreased. In particular, the proportion of expanded molecules with very large expansions (more than 4000 G4C2 repeats) increased significantly in presymptomatic and FTD expansion carriers (p < 1.10^e-4^ and p = 0.006 respectively; Supplementary Table 2). These repeats were also detected in some ALS patients, but the proportion of these large expansions in repeat profiles was globally lower than in other conditions, and independent from the age at sampling (p = 0.98). Although FTD and ALS cohorts were matched for the age at sampling, we nevertheless focused on aged repeat expansion carriers (sampled at > 69y) to control for potential biases between the two conditions (ALS patients being usually described to die earlier than FTD cases, that could explain the lower proportion of very larger expansions in aged carriers). In this small subset of carriers including nine patients with FTD and seven with ALS plus one presymptomatic carrier, the same trends as those detected in the whole cohort were observed (Supplementary Figure 5). In the oldest ALS patients, the proportion of expansion > 4000 G4C2 repeats remained low (< 6 %), whereas this proportion was higher in the asymptomatic carrier and FTD-age matched patients (from 5 to 27%). Although differences were not statistically significant due to the small sample size of this subset, these analyses confirmed previous observations, excluding biases due to different ages at sampling. Interestingly, trends in correlations were very close between presymptomatic carriers and FTD patients for almost all intervals of repeat numbers, in lines with global correlations described in Figure 5. This reinforces the similarity of correlation patterns between presymptomatic and FTD, distinct of ALS expansion carriers.

**Figure 6.**
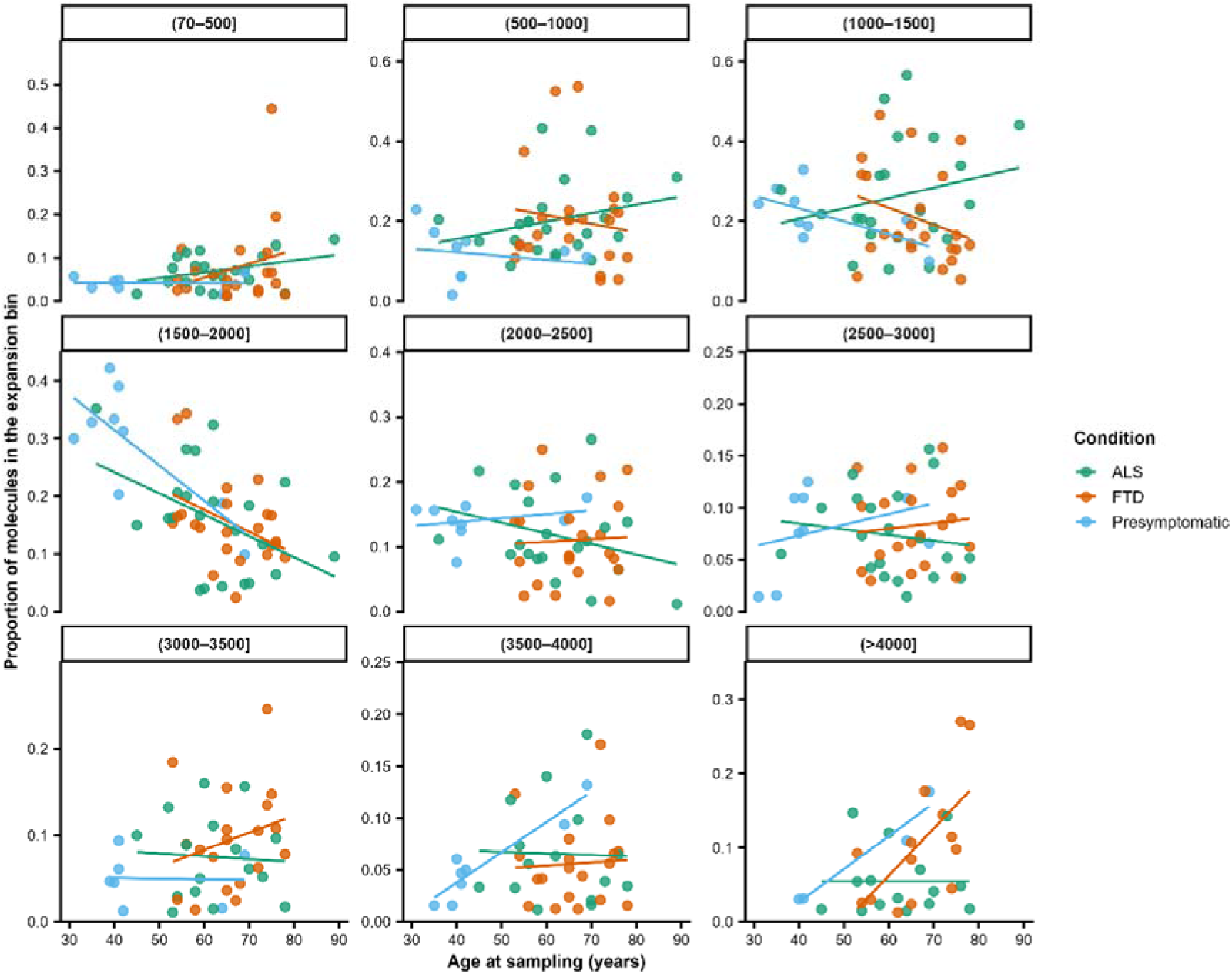
Proportion of G4C2 repeat subclasses as a function of age at sampling. Each panel shows the scatterplot correlation between the proportion of molecules within a specific repeat size bin (bin width of 500 repeats, from (70–500] to (>4,000]) and age at sampling in ALS (green), FTD (orange), and presymptomatic carriers (blue). Individual data points represent individual subjects, shown alongside fitted linear regression lines.

To summarize, the analysis of repeat profiles revealed alternate effects of the age at sampling in carriers according to the clinical condition. A slight enrichment corresponding to the most abundant repeat class (∼mode of repeat profiles) was observed in patients with ALS, whereas we observed a greater repat instability in favor of larger expansions in asymptomatic and FTD expansion carriers along with older ages at sampling. Altogether, these analyses illuminated highly distinct dynamics of repeat instability across clinical conditions.

## Discussion

Since the identification of the pathogenic G4C2 expansion in the first intron of *C9orf72* gene as a major cause of genetic ALS and FTD, the role of expansion size variability in clinical heterogeneity has remained unclear. Here we analyzed repeat profiles between asymptomatic, ALS and FTD expansion carriers using OGM. This method, based on a comparison of optical maps from a sample with a reference map, is particularly well suited for the detection of structural variants, including large expansions such as the *C9orf72* expansion^14^. We first validated this method by comparing it to existing approaches commonly used estimate *C9orf72* repeat numbers. Comparison of classical expansion metrics obtained by OGM with those provided by Southern blotting yielded highly concordant results in the estimation of classical metrics such as mean or median G4C2 repeat numbers. Although correlations with long-read sequencing (LRS) were also significant, this comparison revealed a global underestimation of mean repeat number by LRS, a phenomenon previously reported in multiple independent studies^11–13^. Finally, the allelic balance between molecules carrying the pathogenic expansion and those carrying the normal allele reached the expected 50% threshold in heterozygous carriers, whereas LRS is also known to preferentially sequence the normal allele. Altogether, these technical comparisons showed that OGM is valuable not only to estimate repeat number, but also for providing a more comprehensive view of the allelic distribution allowing a more precise analysis of expansion mosaicism within a sample.

We took advantage of this high level of precision to apply OGM on a cohort of *C9orf72* repeat expansion carriers. Analysis of the whole cohort revealed a marked inter-individual variability in these profiles. Unsupervised hierarchical clustering of these profiles identified distinct groups of individuals based on similarities in distribution shapes. Considering solely repeat profiles obtained from OGM in blood using allelic distribution metrics failed to predict clinical phenotype, consistent with previous Southern-blot or LRS-based studies. However, we identified a small subgroup enriched for patients with a majority of FTD cases underlying a distinct distribution of repeat profiles. The fact that this unsupervised clustering revealed similarities and differences between expansion profiles suggests the existence of non-random factors influencing the allelic distribution of expanded alleles.

In this work, we detected discrepancies of repeat profiles in blood according to the age at sampling between ALS and FTD repeat expansion carriers. The size of this expansion was positively correlated with the age at sampling in FTD cases while it slightly decreased in ALS cases. Differential correlations with the age at sampling between ALS and FTD phenotypes were principally observed for the two tails of the distribution, suggesting different dynamics of contractions vs. expansions in the two clinical groups. In summary, ALS patients showed an age-related increase of repeats with [500-1500] G4C2. Conversely, in FTD patients, the proportion of [500-1500] repeats decreased while the proportion of larger expansions (≥ 2000 G4C2 unit) increased, especially for expansions with more than 4000 repeats. This age-related effect on repeat numbers strongly suggests a different – and almost opposite - dynamics of somatic variability in ALS and FTD patients. This hypothesis was supported by the analysis of the instability index which was also increasing in an age-dependent manner in FTD patients but not in ALS. A classical comment is that ALS patients may die before patients with FTD. However, here the two cohorts were matched for the age at sampling (p = 0.21). In addition, we performed a supplementary analysis in the subgroup of the oldest *C9orf72* carriers, sampled after 69 years of age. In these 16 patients, the median size of the expansion was higher in FTD patients versus ALS (n = 9 and n = 7 respectively), and the proportion of very large repeats (>4000 G4C2) was also higher in FTD patients versus age-matched ALS patients (Supplementary Figure 5). These differences were non-significant (data not shown), probably due to the small number of individuals in this age group, but they are in line with the trends observed in the whole cohort, arguing against the fact that FTD patients arguing against the hypothesis that these large expansions do not appear in ALS patients because they could have died earlier than FTD patients. As expected, the disease duration was significantly shorter in the ALS group which could also appear as an argument to explain the absence of large repeats, if they were linked to the onset of the disease. However, the analysis of presymptomatic carriers argued against that point. Indeed, we observed similar age-dependent correlations on expansion profiles between presymptomatic carriers and FTD patients, by contrast to ALS cases. Here the positive correlation between the repeat number and the age at sampling also observed in presymptomatic carriers cannot be explained by disease duration. Altogether, these results suggest that the dynamics of *C9orf72* somatic expansion size changes appear to be distinctive in ALS patients compared to other clinical status. Alternatively, these variations could be a consequence of ALS which could induce specific pathways in blood cells that ultimately affect somatic instability. Although we do not address this question in the present work, our results nonetheless highlight the importance of identifying factors influencing somatic variability, that may ultimately affect clinical expression. This hypothesis therefore remains speculative, as it is based on a cohort of patients with variable ages at sampling rather than on longitudinal sampling from the same carriers. To formally confirm this important point, it would be necessary to follow expansion profiles in carriers over years and correlate these changes with the clinical phenotype.

Here, we chose a design of inclusions with a pure clinical diagnosis (FTD or ALS) to observe changes linked to traits as clearly as possible, if so. However, we cannot exclude that FTD patients enrolled at the time of this study will develop ALS, and inversely, limiting our conclusions on the clinical phenotype to the first predominant symptoms developed by *C9orf72* expansion carriers. Thus, this study warrants independent confirmation using OGM or techniques equally capable of reliably detecting and quantifying very large G4C2 expansions. In addition, the sequence of the expansion itself cannot be analyzed by OGM. So possible variations of sequence changes or methylation influencing the phenotype were not considered in this work. However, to our knowledge, this is the first time that such detailed variations of blood repeat profiles and associated repeat instability are associated with the clinical variability in the *C9orf72* disease.

The influence of repeat number on the clinical expression has already been shown for some other STR loci such as *ATXN1*, *ATXN2* (intermediate alleles being associated with an increased risk to develop ALS vs. pathogenic expansions associated with spinocerebellar ataxia), *ATXN7* (very large repeat associated with congestive heart failure vs. neurological dysfunction for smaller pathological expansions) and *FMR1* (premature ovarian failure for premutations vs. Fragile X syndrome for very large expansions)^16–18^. These examples illustrate how repeat length influences the topography of affected tissues, giving rise to completely different clinical entities. In addition, somatic expansions are also well-known modifiers of the phenotype in expansion diseases like in Huntington’s disease^19,20^. Thus, it is reasonable to speculate that in the *C9orf72* disease, such somatic variations of repeat profiles might influence the evolution of the disease. Interestingly, Somatic instability in brain tissue has already been suggested as *C9orf72* disease-modifiers^6,21–23^. More recently, the age-dependent somatic instability in brain tissue as well as the chromosomal fragility at the *C9orf72* locus has been demonstrated in mouse model^24^. Previous Southern blot-based studies showed that repeat sizes measured in blood were not correlated with those measured in different brain regions^4,6,22,25^. Differences in expansion profiles between ALS and FTD patients may be tissue-specific and limited to blood or only partially reflect processes occurring in the brain, warranting similar approaches in cerebral tissues.

The effect of G4C2 repeat number has been shown to act on several biological hallmarks of the disease. Dipeptides produced by RAN translation of the repeat were shown to be a crucial element that explains the pathogenicity of the expansion^26^. Larger repeats were notably associated with lower levels of dipeptides (poly(GP) and poly(GA)), lower expression of *C9orf72* transcript (intron 1b) in the cerebellum, and a higher proportion of methylated CpG at the *C9orf72* locus in blood^11,12^.Variable levels of DNA methylation at the *C9orf72* locus, linked to the repeat number and age of carriers, were also evoked as a disease modifier^27^. Thus, somatic variations of repeat numbers at the two tails of the allelic distribution (i.e shortest vs. largest expansions) may have the greatest biological impact.

In conclusion, our work sheds new light on distinct patterns of somatic instability, enabled by new methodological approaches. Previous LRS-based studies have already provided important advances and precisions compared with Southern blot studies, illustrating the value of new techniques to revisit expansion size / phenotype relationships.

How these somatic variations influence, and/or could serve as biomarkers of disease progression is also a key question in the context of therapeutic trials that could target somatic G4C2 expansions in carriers as in some other expansion diseases. Finally, investigating the somatic variability of the pathogenic repeat, and deciphering the factors influencing this instability may be more relevant to better understand the role of this expansion in the *C9orf72* in clinical variability.

## The PREV-DEMALS/STRATALS study group include

Philippe Couratier (CHU Limoges), Giorgia Querin (ICM, Paris), Pierre-François Pradat (LIB, APHP), Florence Pasquier (CHU Roger Salengro, Lille), Thibaud Lebouvier (CHU Roger Salengro, Lille), Isabelle Le Ber (Hôpital Pitié-Salpêtrière, Paris), Veronique Marchand-Pauvert (LIB), Aleksandra Lakmy (LIB), Armelle Rametti-Lacroux (ICM), Daisy Rinaldi (Hôpital Pitié-Salpêtrière, Paris), Adeline Rollin-Sillaire (CHU Roger Salengro, Lille), and David Wallon (CHU Charles Nicolle Rouen).

## AKNOWLEDGMENTS

We are very grateful to patients and their families who were enrolled in this study. art of this work was carried out in the DNA and cell bank core facility of the Paris Brain institute. We also thank the data analysis core facility (DAC) from the Paris Brain Institute for their technical and scientific assistance. We also thank the Genomics Core Facility, Department of Translational Research (Institut Curie, Paris, France), for their technical assistance to perform the sizing of the G4C2 repeat number using optical genome mapping.

## DATA AVAILABILITY

Custom scripts and code to estimate the size (number of G4C2 repeat units) in *C9orf72* from Bionano Genomics Optical Genome Mapping (OGM) single-molecule alignments are publicly available in a GitHub repository https://github.com/Genosfert/ogm-tandem-repeat-sizing under the MIT license.

## Supplementary information

**Supplementary Figure 1.**
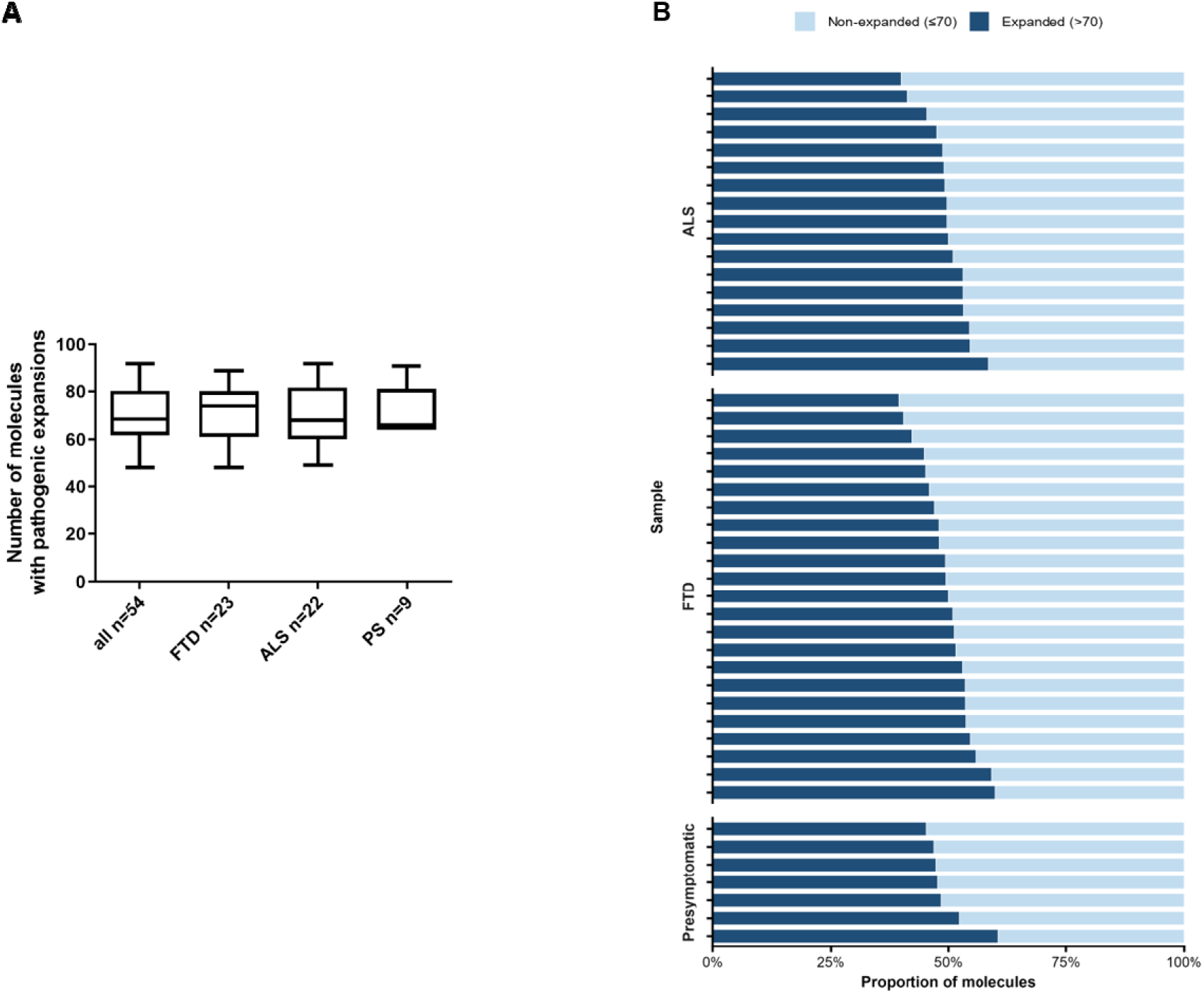
Detection of *C9orf72* expanded alleles using Optical Genome Mapping. **A)** Box plots representing the number of molecules carrying the pathological G4C2 expansion in the whole cohort and in sub-groups. FTD: Frontotemporal Dementia; ALS: Amyotrophic Lateral Sclerosis; PS: Presymptomatic. Whiskers indicate mon and max, , Boxes represent Q1, Q3 and median. **B)** Ratio (%) of expanded / non-expanded alleles according to clinical sub-groups.

**Supplementary Figure 2.**
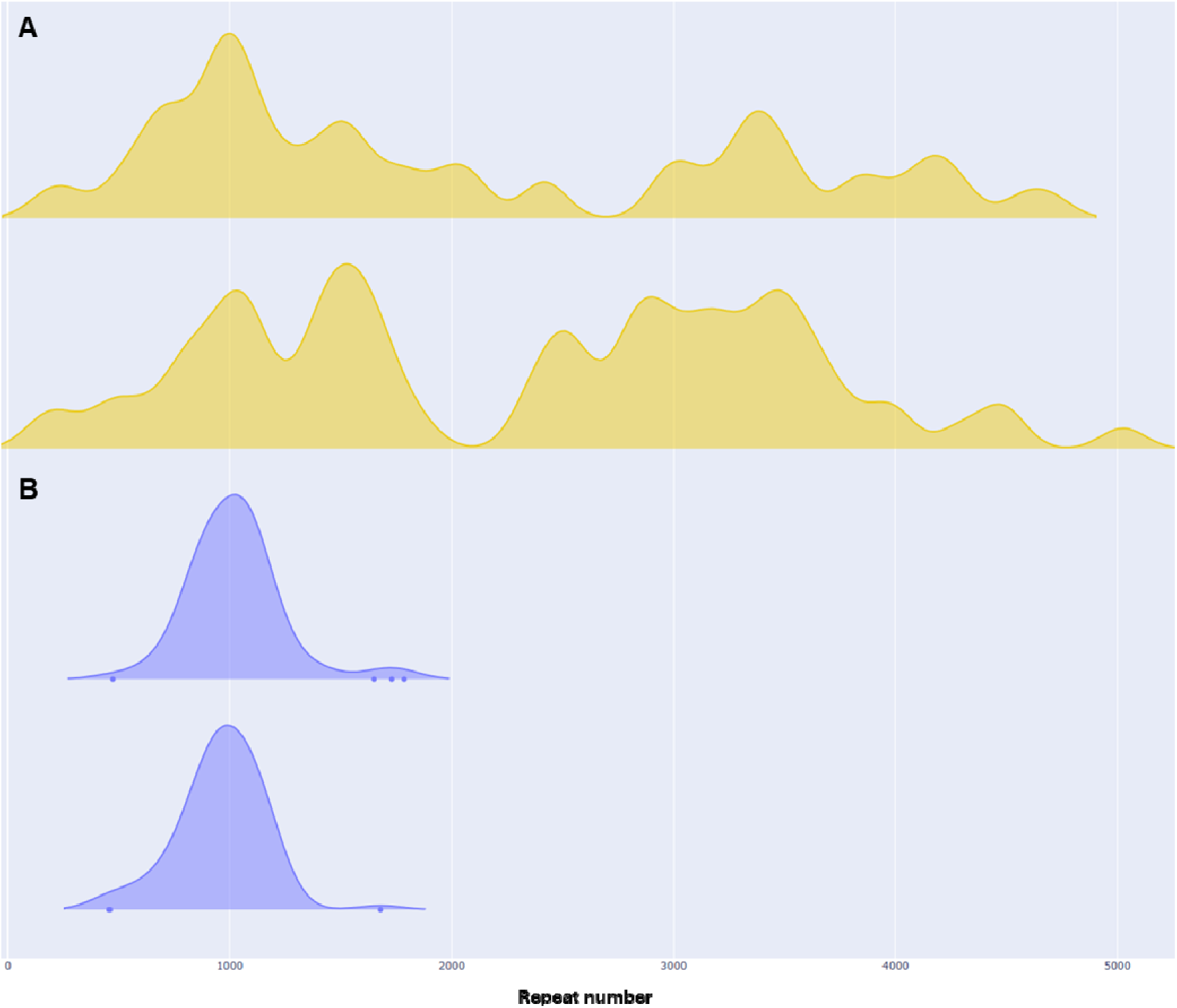
*C9orf72* Allelic distribution (expanded alleles, density curves) from OGM in two patients sampled twice. **A)** Patient with FTD at sampling, six month-interval, Top = first sampling; **B)** Patient with ALS at sampling, one year-interval, Top = first sampling.

**Supplementary Figure 3.**
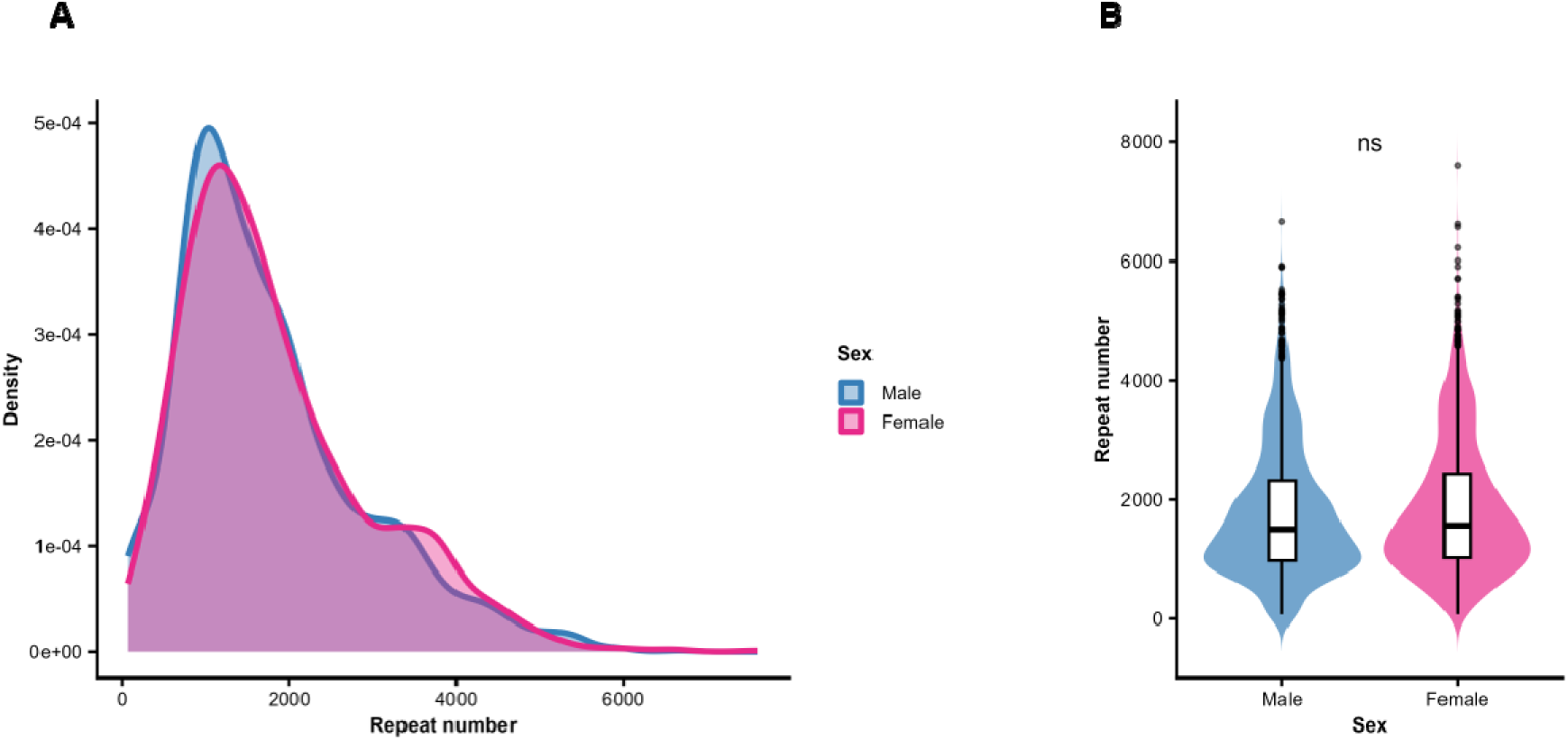
Alllelic distributions of the *C9orf72* expansion in carriers (expanded alleles) according to sex. **A)** Ridgeline plots of the consensus distributions in Females and Males. **B)** Violin plots of allelic distributons.

**Supplementary Figure 4.**
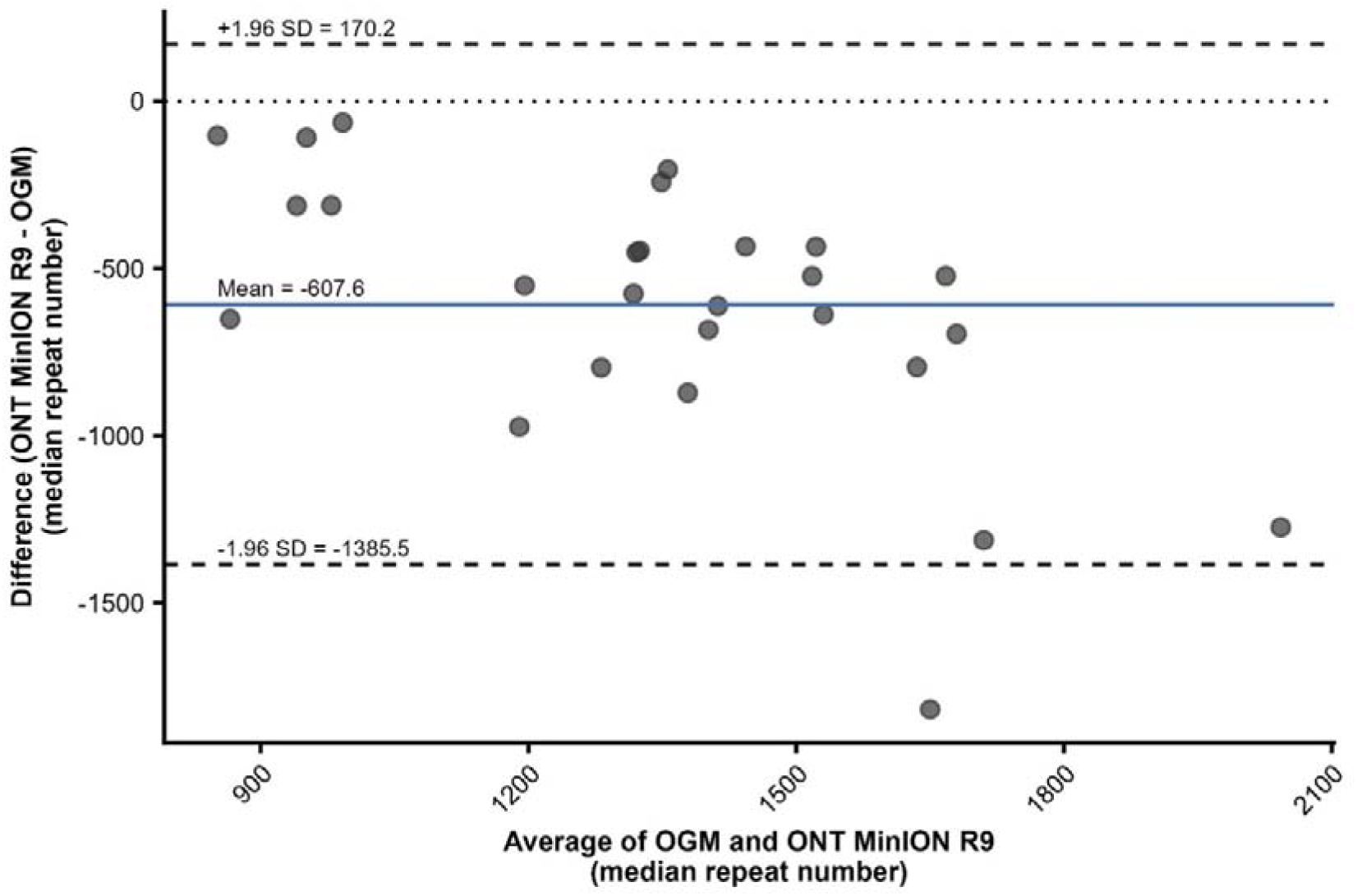
Bland-Altman plot testing the agreement between OGM and ONT-LRS to estimate G4C2 median repeat number.

**Supplementary Figure 5.**
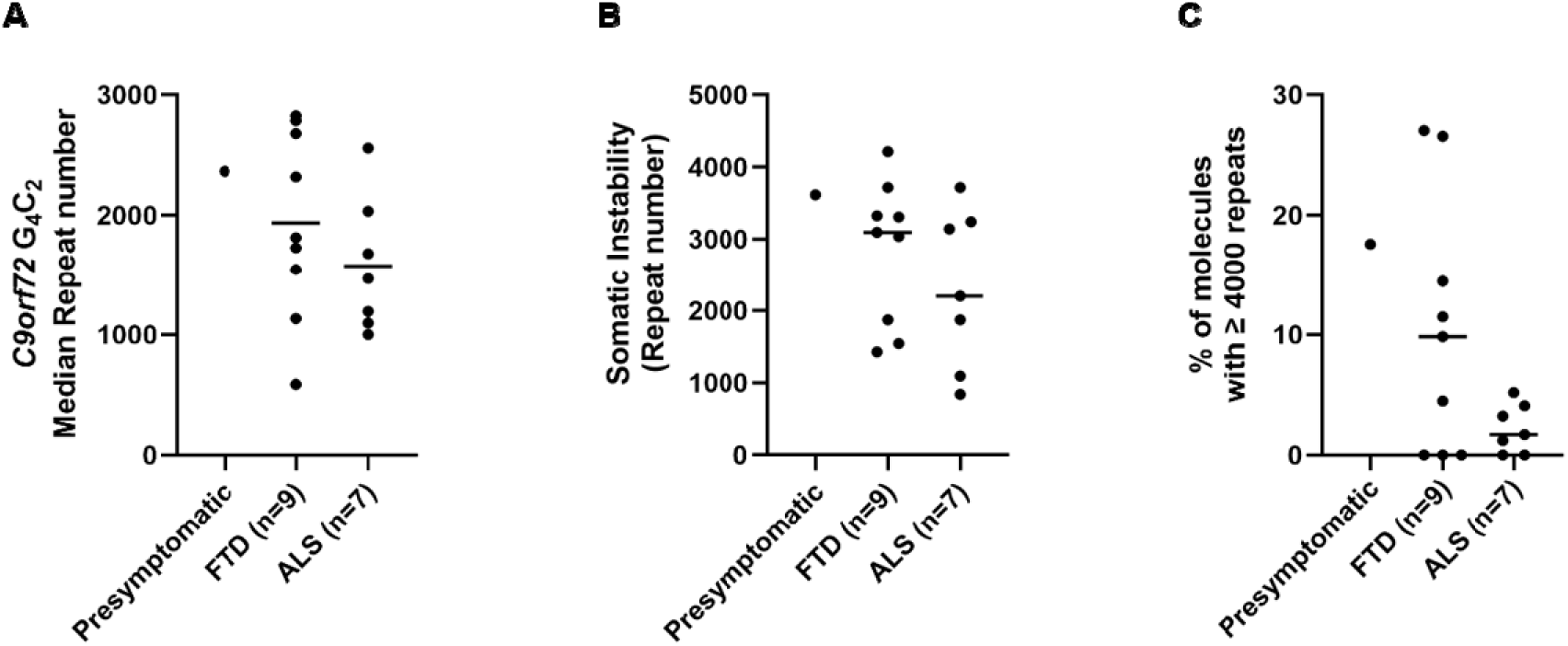
Sizing of the *C9orf72* repeat expansion by Optical Genome Mapping in carriers sampled at > 69 years of age. Median repeat number (**A**) and Somatic instability (**B**) in presymptomatic, FTD and ALS patients. **C**) Proportion (%) of expansions with more than 4000 G4C2 repeats in presymptomatic, FTD and ALS patients. Each point represents an expansion carrier.

**Supplementary Table 1.** Principal metrics of the *C9orf72* G4C2 expansion in each cluster obtained from the unsupervised hierarchical clustering of allelic distributions. Repeat numbers are reported as median (Q1, Q3).

|  | Cluster 1 | Cluster 2 | Cluster 3 |
| --- | --- | --- | --- |
| n | 23 | 20 | 11 |
| GGGGCC repeat number |  |  |  |
| Mean | 1431 (1110, 1854) | 1913 (1663, 2110) | 2500 (2033, 2742) |
| Median | 1415 (1024, 1605) | 1676 (1539, 1909) | 2524 (1882, 2680) |
| Standard Deviation | 753.8 (561.6, 974.2) | 989 (839.1, 1225) | 1319 (901.3, 1375) |
| Somatic Instability index | 1857 (1210, 2381) | 2487 (2030, 3231) | 3309 (2203, 3501) |

**Supplementary Table 2.** Correlations between *C9orf72* G4C2 repeat number and age at sampling for each interval (bin = 500), according to the clinical condition. Spearman’s rho and associated p-values are indicated.

| bin | Condition | n | rho | p value |
| --- | --- | --- | --- | --- |
| (70–500] | ALS | 20 | 0,23 | 0,33 |
| (70–500] | FTD | 21 | 0,16 | 0,49 |
| (70–500] | Presymptomatic | 7 | -0,01 | 0,98 |
| (500–1000] | ALS | 22 | 0,26 | 0,24 |
| (500–1000] | FTD | 23 | -0,05 | 0,83 |
| (500–1000] | Presymptomatic | 9 | -0,34 | 0,37 |
| (1000–1500] | ALS | 22 | 0,20 | 0,36 |
| (1000–1500] | FTD | 23 | -0,32 | 0,14 |
| (1000–1500] | Presymptomatic | 9 | -0,59 | 0,09 |
| (1500–2000] | ALS | 22 | -0,38 | 0,08 |
| (1500–2000] | FTD | 23 | -0,42 | 0,05 |
| (1500–2000] | Presymptomatic | 9 | -0,56 | 0,12 |
| (2000–2500] | ALS | 20 | -0,25 | 0,28 |
| (2000–2500] | FTD | 22 | 0,09 | 0,70 |
| (2000–2500] | Presymptomatic | 9 | 0,20 | 0,60 |
| (2500–3000] | ALS | 20 | -0,27 | 0,25 |
| (2500–3000] | FTD | 20 | 0,09 | 0,70 |
| (2500–3000] | Presymptomatic | 9 | 0,49 | 0,18 |
| (3000–3500] | ALS | 16 | -0,03 | 0,91 |
| (3000–3500] | FTD | 19 | 0,27 | 0,26 |
| (3000–3500] | Presymptomatic | 7 | -0,04 | 0,94 |
| (3500–4000] | ALS | 15 | -0,07 | 0,80 |
| (3500–4000] | FTD | 19 | 0,12 | 0,62 |
| (3500–4000] | Presymptomatic | 8 | 0,84 | 0,01 |
| (>4000] | ALS | 15 | 0,01 | 0,98 |
| (>4000] | FTD | 14 | 0,69 | 0,01 |
| (>4000] | Presymptomatic | 4 | 1 | <10<sup>e-4</sup> |

